# The effect of enzyme and protein containing toothpaste on gingival condition: a randomised controlled study

**DOI:** 10.1101/2025.05.07.25326607

**Authors:** Xiao Hu, PingNeng Zhang, Lu Zhang, Jane R Matheson, Shengnan Lin, Jianing N. Sun, Cristina Delfanti, Jiangang Tian, Ashok K Gupta, Ruizhe Huang

## Abstract

**Background:** Accumulation of dental plaque can lead to gum problems. A fluoride toothpaste containing naturally occurring enzymes and proteins has been shown to improve gingival health and reduce supragingival plaque in European populations. The objective of the current study was to evaluate the ability of this toothpaste to improve gingival condition and reduce supragingival plaque in an alternative study population.

**Methods:** This was a double-blind (participant, examiner, investigator, statistician), randomised, parallel group, efficacy study conducted at a Chinese university dental hospital. Participants (age 18-70) with a mean gingival index (GI) ≥ 1.0 and mean modified Quigley and Hein plaque index (MQHPI) ≥ 1.5 were randomised using sex stratification to twice-daily brushing with either test toothpaste (n=127) or control fluoride toothpaste (n=124) for 26 weeks. Assessments of GI (primary outcome) and MQHPI were conducted at baseline and after 4, 13 and 26 weeks of product use. Results were analysed using ANCOVA model for between product comparison (α= 0.05).

**Results:** One hundred and six participants using test toothpaste and 92 using control toothpaste completed the study. The test toothpaste was more effective than control toothpaste in improving gingival condition after 4, 13 and 26 weeks of product use, with estimate product differences in GI of -0.18 at week 4 (p < 0.0001, 95% CI -0.23 to -0.12), -0.16 at week 13 (p < 0.0001, 95% CI -0.20 to -0.11) and -0.53 at week 26 (p < 0.0001, 95% CI -0.60 to -0.46). The test toothpaste was also significantly more effective than control toothpaste in reducing supragingival plaque after 4, 13 and 26 weeks of product use, with estimated product differences of -0.29 at week 4 (p < 0.0001, 95% CI -0.38 to -0.21), -0.35 at week 13 (p < 0.0001, 95% CI -0.45 to 0.25) and -0.96 at week 26 (p < 0.0001, 95% CI -1.06 to -0.85).

**Conclusion:** The study demonstrated that a toothpaste containing naturally occurring enzymes and proteins significantly improved gingival health and reduced supragingival plaque compared to a control toothpaste, after 4, 13, and 26 weeks of use.

## Background

Gingivitis is an acute or chronic inflammation of the gingival tissue surrounding the tooth, characterized by redness, swelling and bleeding of the gums [1]. It is a public oral health problem which is highly prevalent across different populations. In China, the fourth national oral health epidemiology survey conducted in 2015 demonstrated that 87.4% of 35- to 44-year-olds had gum bleeding, an increase of 10.1% compared to 2005 [2]. In the United Kingdom, 52.9% of adults were shown to have gingival bleeding in the Adult Dental Health Survey of 2018 [3]. In separate studies conducted in 2017, 76% of UK dentist attendees, aged 18-92, had bleeding on probing [4] while in a French study 63.2% of adults self-reported gum bleeding [5].

Dental plaque is a primary cause of gingivitis [6]. The bacteria in plaque produce enzymes and toxins that irritate soft tissues surrounding the teeth (the gingiva), leading to gum inflammation and weakening of the connection between the tooth and gum [7]. If gingivitis is not effectively managed, it can progress to periodontitis, negatively impacting quality of life and having systemic consequences [8,9,10,11]. Adequate oral hygiene practices play a crucial role in preventing and controlling gingivitis by reducing the accumulation of dental plaque [8].

Twice-daily brushing with toothpaste serves to physically remove plaque, whereas incorporating antimicrobial agents into toothpaste can enhance plaque control [12,13]. A fluoride toothpaste incorporating a three-enzyme system (amyloglucosidase, glucose oxidase and lactoperoxidase) has been developed to enhance natural microbial control in the oral cavity. Amyloglucosidase and glucose oxidase generate hydrogen peroxide from polyglucans, and lactoperoxidase oxidizes salivary thiocyanate into hypothiocyanite in the presence of hydrogen peroxide [14,15]. Both hydrogen peroxide and hypothiocyanite exhibit potent antimicrobial activity and have been shown to effectively inhibit periodontal pathogens [14,16,17]. This toothpaste also contains three proteins reflecting saliva’s natural defence mechanisms: bovine colostrum containing immunoglobulin IgG, lactoferrin and lysozyme. Immunoglobulin provides protection against infection, lactoferrin inhibits the metabolic activity of several oral pathogens and lysozyme breaks down peptidoglycan, which is an essential part of the cell wall of the gram-positive bacteria [17,18,19,20].

With regular use, this toothpaste containing enzymes and proteins has been shown to promote an overall shift in the bacterial community, with an increase in gum health-associated bacterial species and a decrease in abundance of periodontitis-associated bacterial species [16]. Furthermore, there is evidence that toothpaste containing enzymes and proteins enhances salivary defences by increasing the levels of antimicrobial compounds lysozyme and hydrogen peroxide *in vivo*, as well as hypothiocyanite *in vitro* [21]. A clinical study in a United Kingdom (UK) population demonstrated significant improvements in gum health after 13 weeks use of the toothpaste containing enzymes and proteins compared to control fluoride toothpaste [15]. Furthermore, an epidemiological study in Denmark reported better gum condition in participants who had used the toothpaste containing enzymes and proteins for at least 12 months compared to control group who used toothpastes that did not contain antimicrobial actives [22]. The purpose of this current clinical study was to investigate if the gum health benefits provided by this toothpaste were transferrable to another study population.

The primary objective of the current study was to compare the efficacy of this enzyme and protein containing toothpaste with a regular fluoride toothpaste in Chinese individuals with gum problems after 13 weeks of twice-daily use, as measured by gingival index (GI) [23]. Secondary objectives included the efficacy as measured by GI after 4- and 26-weeks’ use, and plaque reduction measured by modified Quigley and Hein plaque index (MQHPI) [24] after 4-, 13- and 26-weeks’ use.

The null hypothesis was that the enzyme and protein containing toothpaste would not improve gingival condition nor reduce supragingival plaque compared to regular fluoride toothpaste in Chinese individuals with gum problems after 13 weeks of twice-daily use.

## Materials and Methods

### Participants and study design

This was a 26 week, double-blind (participant/examiner/statistician/investigator), randomised, two group, parallel design study conducted at a University Dental Hospital in Xi’an, China from October 2018 to April 2019. The overall test design, as well as the measurement of gingival health, followed the guideline WS/T 326.3-2010 for efficacy evaluation on inhibiting dental plaque and/or reducing gingival inflammation [25]. The study was approved by a local ethics committee prior to enrolment of the first participant. This study is reported according to the Consolidated Standards of Reporting Trials statement (CONSORT 2010) for parallel group randomised trials [26].

Volunteers attending screening were informed of the study purpose, and those who gave written informed consent were assessed for eligibility according to inclusion and exclusion criteria and had a dental assessment. The key inclusion criteria were: 1) Aged 18-70 years, of either sex and in good general health; 2) Having at least 20 natural teeth without subgingival calculus including 5 teeth (excluding 3rd molars) in each quadrant, which can be assessed; 3) Having mean Gingival Index (GI) ≥ 1.0 and mean Modified Quigley and Hein Plaque Index (MQHPI) ≥ 1.5 at screening and baseline. Participants who had the following situations were excluded: 1) Pregnant or breast feeding mothers; 2) Having obvious signs of untreated caries or significant periodontal disease; 3) Full or partial dentures wearers; 4) Current orthodontic treatment; 5) Smokers or those who have a recent smoking history, including e-cigarettes; 6) Diabetics; 7) Having a course of anti-inflammatory, antimicrobial or statin drugs within 4 weeks of screening.

### Study procedure

Qualified participants were randomly allocated to use test or control toothpaste (Table 1) by study staff. The examiner was blinded to product allocation. Stratified block randomisation schedule (block size four) was generated by the statistician using Proc Plan procedure in SAS 9.4 (SAS Institute Inc., Cary, NC, USA) suitable for 2-group parallel design. Sex was considered as a stratification factor. Test toothpaste was provided in a plain white tube while control toothpaste was overwrapped in white to aid study blinding. The labels on both toothpastes differed only by an alphanumeric code. Additionally, to maintain uniformity between groups, all participants were provided with a soft bristle toothbrush (Sanxiao, China).

**Table 1.** Study Products.

| Toothpaste | Description |
| --- | --- |
| Test toothpaste | Unilever Enzymatic technology toothpaste - 1450 ppm Sodium fluoride, lactoferrin, colostrum, amyloglucosidase, glucose oxidase, lactoperoxidase and potassium thiocyanate |
| Control toothpaste | Sensodyne Pronamel® - 1450 ppm Sodium fluoride |

During the study phase, participants were asked to only use their allocated toothbrush and toothpaste to brush for at least one minute twice a day, in the morning and at night, for 26 weeks. The amount of paste used each time should cover the bristle head of toothbrush. Participants were educated with the correct brushing method and brushed teeth on site at baseline, week 4 and week 13 visits.

The participants underwent clinical assessments at the study site by a single assessor at baseline, and after 4 weeks, 13 weeks and 26 weeks of product use.

### Assessment of efficacy

Each test session comprised two efficacy measurements. Both measurements were carried out on all scorable teeth except the third molars. Six sites on each tooth were assessed, three on the facial surfaces (buccal, mesial and distal) and three on the palatal/lingual surface (palatal/lingual, mesial and distal).

The gingival condition was assessed using gingival index (GI) [23]. The scoring criteria were: 0 = Absence of inflammation; 1 = Mild inflammation - slight change in colour and little change in texture; 2 = Moderate inflammation – moderate glazing, redness, oedema, and hypertrophy (bleeding on pressure); 3 = Severe inflammation - marked redness and hypertrophy (tendency to spontaneous bleeding).

Supragingival plaque was disclosed by Red-cote and assessed using modified Quigley and Hein plaque index (MQHPI) [24]. The scoring criteria were: 0 = No plaque; 1 = Separate flecks of plaque at the cervical margin of the tooth; 2 = A thin continuous band of plaque (up to 1 mm) at the cervical margin of the tooth; 3 = A band of plaque wider than 1 mm but covering less than 1/3 of the crown of the tooth; 4 = Plaque covering at least 1/3 but less than 2/3 of the crown of the tooth; 5 = Plaque covering 2/3 or more of the crown of the tooth.

For both GI and MQHPI, the whole mouth average per participant was calculated from the sum of all tooth site scores divided by the total number of measured sites.

### Statistical analyses

The sample size was calculated based on a previous clinical study evaluating the same test toothpaste [15]. Assuming a standard deviation of 0.5 for GI, a sample size of 100 participants per group was deemed sufficient to detect a mean difference of 0.2 units on GI between the test and control groups, with a power of 80% and a significance level of 0.05.

As defined in protocol, all randomised participants who had at least one post-baseline efficacy assessment were included in the statistical analysis. Statistical analysis was performed using SAS 9.4 (SAS Institute Inc., Cary, NC, USA). ANCOVA model, with raw values as response, product and sex as fixed effects and baseline as covariate, was conducted for between toothpaste comparisons. The interaction effect for product and sex was significant for MQHPI, hence retained in the final model along with the fixed effects. However, no significant effect was observed for GI, so the interaction was excluded from the final model. Within toothpaste comparisons for each post baseline timepoint were tested using paired t-test.

A post-hoc analysis was performed to derive non-bleeding sites from GI data as following: GI Score (0,1) = 0 (non-bleeding site); GI Score (2, 3) = 1 (bleeding site). Percentage of sites with no bleeding at each timepoint was calculated. Participant level data (percentage of site with no bleeding i.e. P) was transformed into logit as following: Logit=log((P+0.05)/(100.05-P)). Analysis of Covariance (ANCOVA) model was performed for product comparison with logit transformed values as response, product and sex as fixed effects and baseline as covariate.

All tests were two-sided and performed at the 5% significance level.

## Results

A total of 251 participants (test 127/control 124) were randomised to study toothpaste with 198 participants (test 106/control 92) completing 26-week product use phase. Fifty-three participants (test 21/control 32) withdrew during the study (Figure 1, Consort Flow Diagram). Sixteen participants’ baseline data (test 5/ control 11) were excluded from the analysis as they had no post baseline efficacy assessment. All participants included in the analysis were analysed according to the group they were originally assigned. There were no product related adverse events reported during the study.

**Figure 1.**
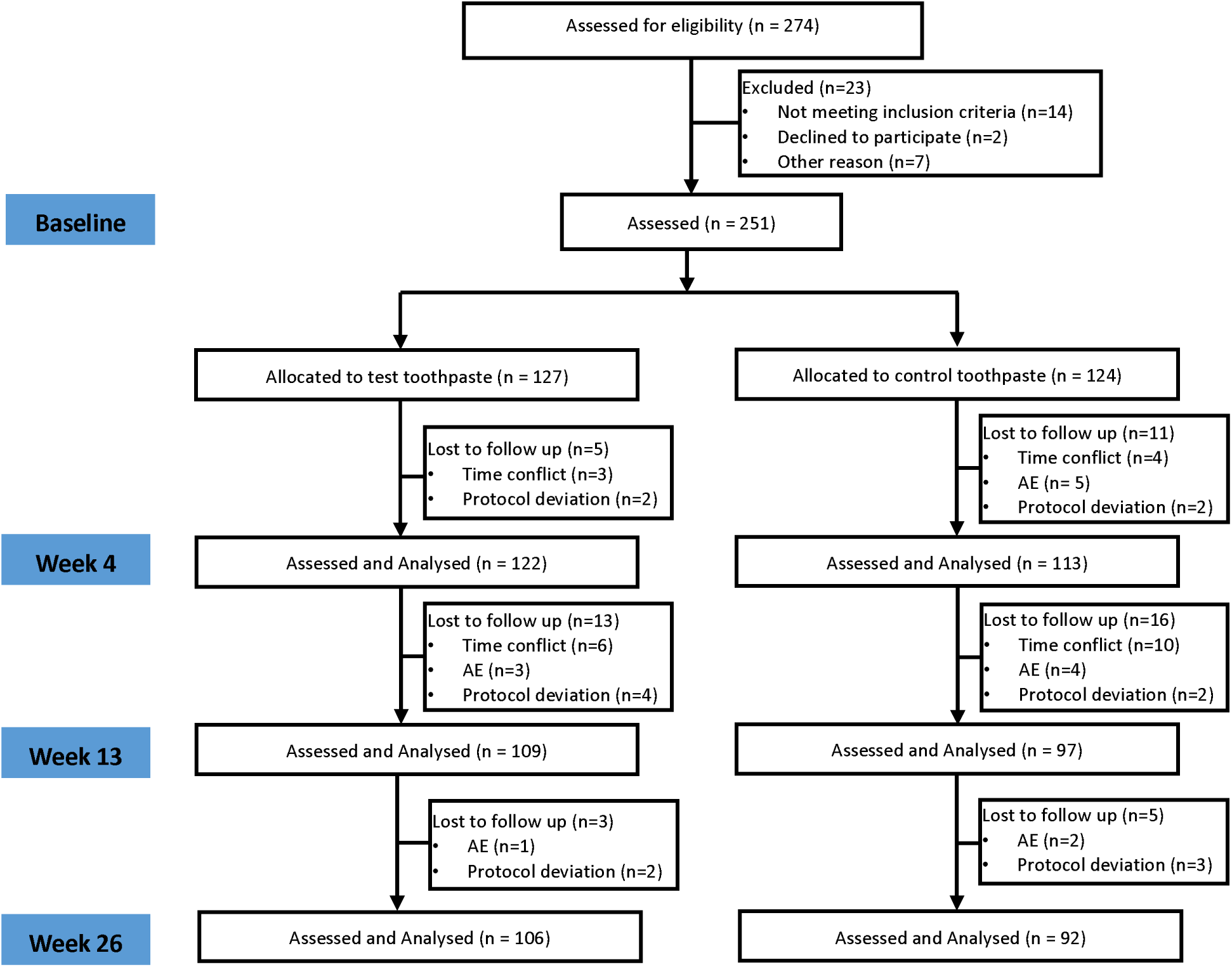
Consolidated standards of reporting trials flow diagram of the study population through the study

The demographic characteristics of randomised study population at baseline are described in the Table 2.

**Table 2.** Summary of Age and Sex for Randomised Participants.

|  | Number of participants |  |  | Age |  |
| --- | --- | --- | --- | --- | --- |
|  | Male | Female | Total | Mean (SD) | Range |
| Test | 40 | 87 | 127 | 40.51 (12.98) | 19-69 |
| Control | 37 | 87 | 124 | 43.02 (12.02) | 21-67 |

Table 3 provides a summary of the unadjusted mean values for GI and MQHPI at baseline, 4-week and 13-week timepoints, plus the baseline adjusted means for the between product comparison at each timepoint. Both test and control toothpastes showed significant reductions in GI and MQHPI compared to baseline, after 4, 13 and 26 weeks of product use (p<0.05 for all, Table 3). Relative to control toothpaste, test toothpaste exhibited a significant reduction in GI after 4, 13 and 26 weeks of product use (p<0.0001 for all, Table 3). There was also a statistically significantly greater reduction in MQHPI for test toothpaste compared with control toothpaste after 4, 13 and 26 weeks of product use (p<0.0001 for all, Table 3).

**Table 3.**
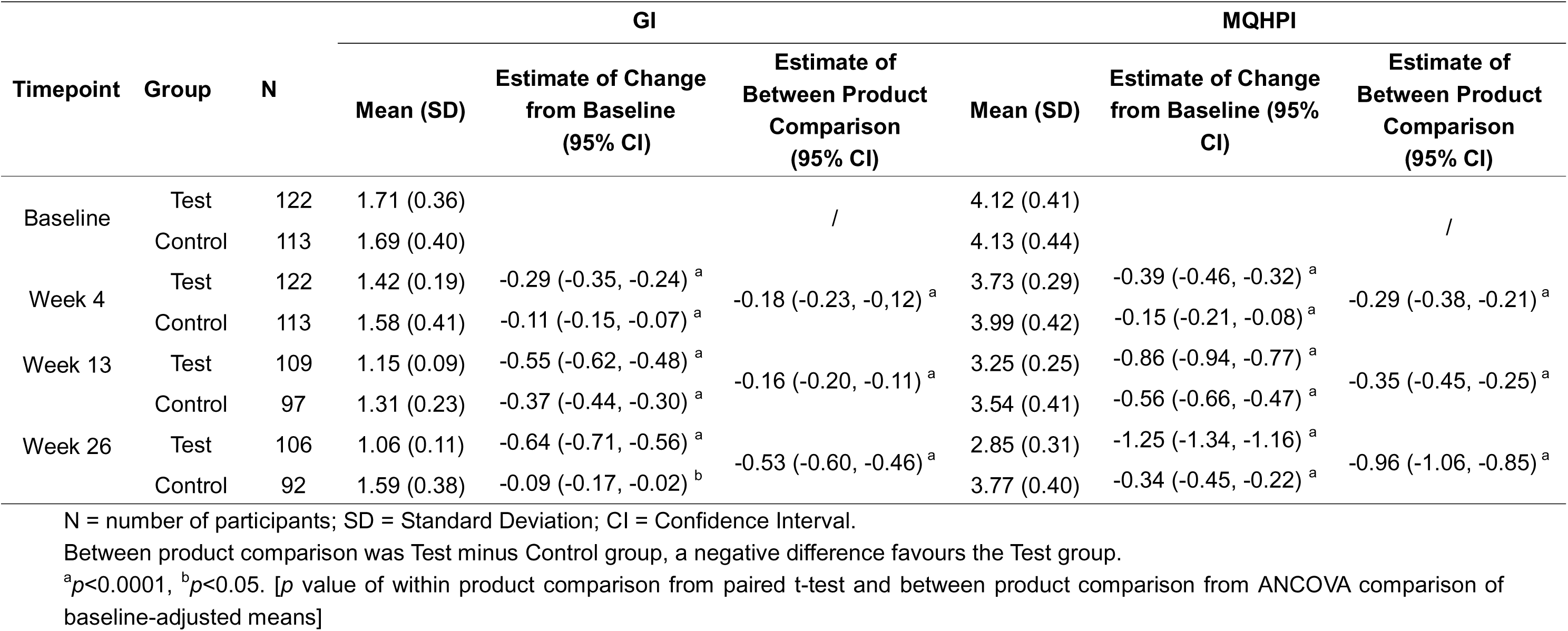
Results on GI and MQHPI assessment.

Figure 2 and Table 4 display the percentage of gum sites that were non-bleeding at each timepoint for both test and control toothpastes. At baseline, participants on the test toothpaste had on average 35.7% non-bleeding sites in the mouth. This increased to 59% after 4 weeks, 84.8% after 13 weeks and 93.8% after 26 weeks. The between product comparison showed that participants on the test toothpaste had significantly higher percentages of non-bleeding sites than control toothpaste at all three timepoints (p<0.01).

**Figure 2.**
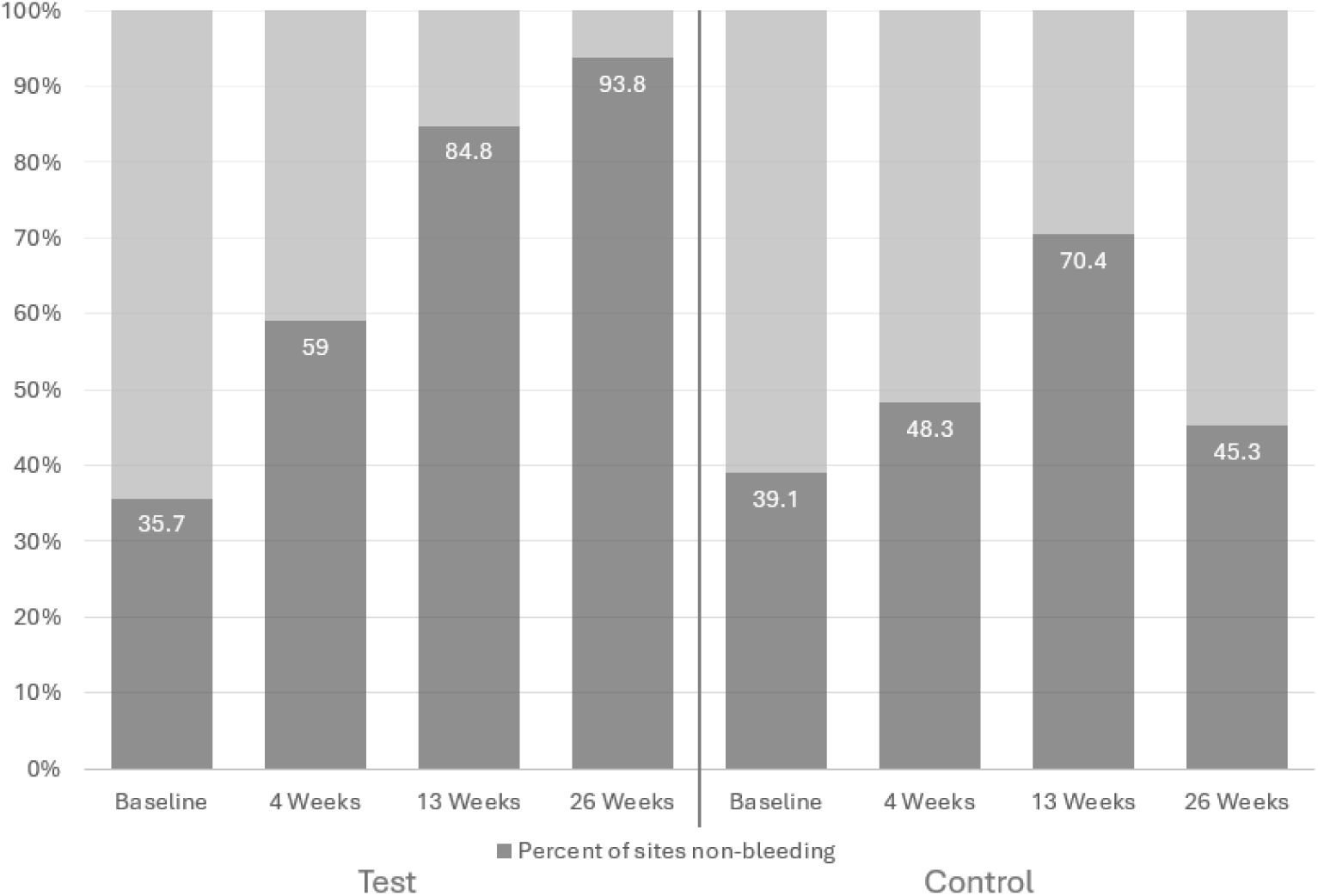
Percentage of gingival sites considered non-bleeding (gingival index score of 0 or 1) at each timepoint. At 6 months 93.8% of sites in the test group were non-bleeding as compared to 45.3% in the control toothpaste group.

**Table 4.**
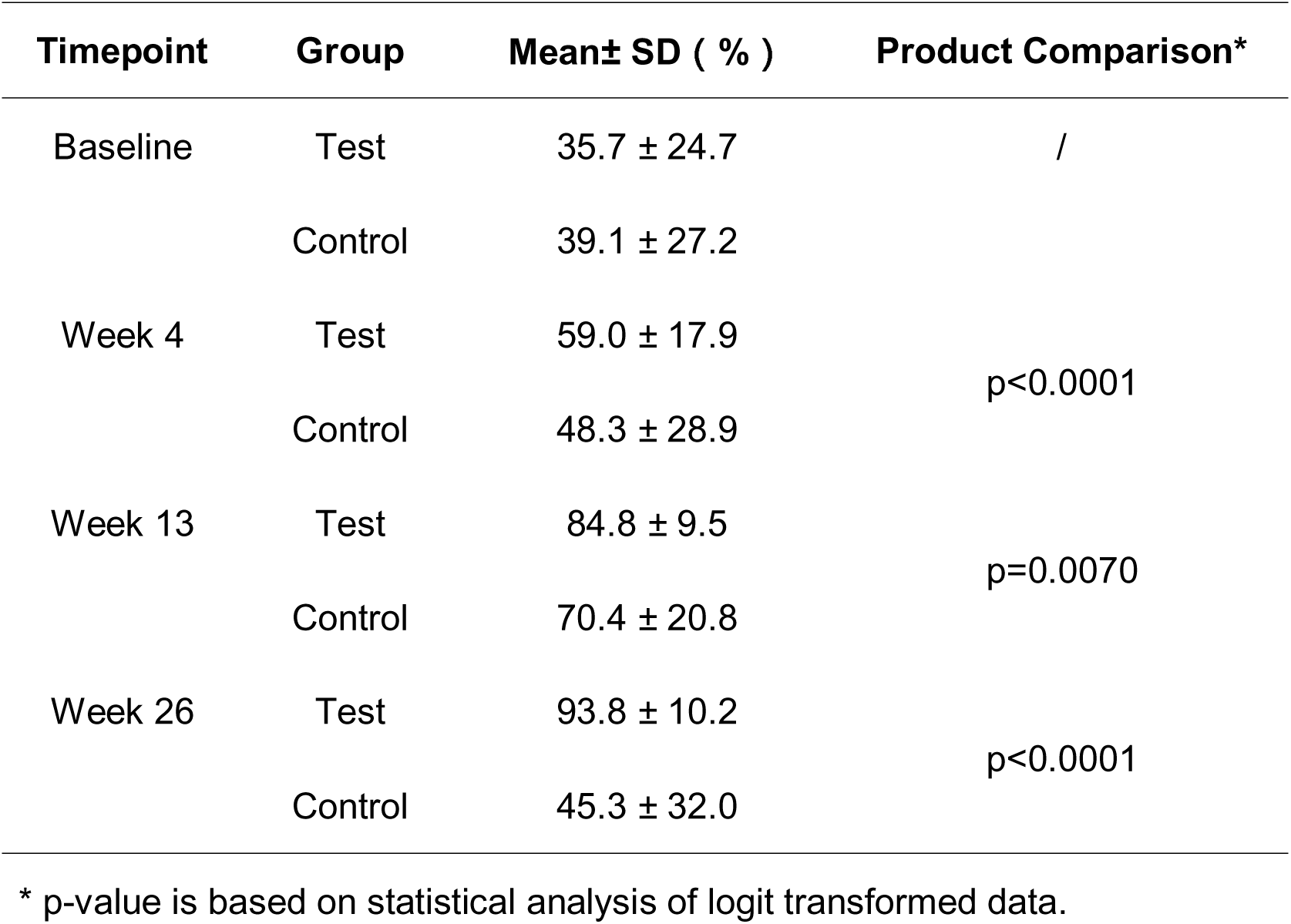
Percentage of non-bleeding sites from GI data.

## Discussion

This double-blind, randomised, clinical study provides further evidence that the enzyme and protein containing toothpaste provides gum health benefits to those who suffer from mild to moderate gum problems. The test toothpaste showed significant improvements in gingival condition and reductions in supragingival plaque from 4 weeks, with benefits increasing over the 26-week time frame of the study. After 6 months of using the test toothpaste twice daily, the percentage of non-bleeding tooth sites (gingival index 0 or 1) increased from 35.7% at baseline to over 93%, which is significantly higher than the 45.3% achieved with the control toothpaste.

It is of note that statistically significant improvements from baseline were also observed with the control toothpaste for both gingival condition and supragingival plaque levels. At each test visit, participants were required to brush teeth on site as per instruction, which likely influenced tooth brushing behaviour, thereby enhancing plaque control and contributing to improvement of gingival condition during participation. It is speculated that the 3-month interval between brushing instruction given at week 13 and the last examination at week 26 reduced the impact of this instruction. Consequently, participants might have returned to their normal tooth brushing behaviour, as shown by the fact the gum condition and plaque levels for the control toothpaste at 26 weeks were worse than those at 13 weeks. Importantly, the effect of the test toothpaste on improving gum health was evident and superior to the negative control, over and above the effect of the brushing instructions.

The gum health efficacy of this enzyme and protein containing toothpaste has previously been evaluated in a double-blind, randomised clinical study in a UK population [15]. The toothpaste also reduced gingival inflammation, gingival bleeding and plaque levels in this study over the 13 week test phase as compared to negative control toothpaste [15]. Additionally, an epidemiological study carried out in Denmark showed lower levels of gingival inflammation, gingival bleeding and plaque in those who had used the toothpaste containing enzymes and proteins for at least 12 months as compared to those who had been using toothpastes without antimicrobial actives for a similar period [22]. Taken together, the results from the double-blind, randomised clinical studies and the real-world epidemiology study provide strong evidence that the gum health benefits of the enzyme and protein containing toothpaste are robust and are generalisable to all individuals who suffer from mild to moderate gum problem.

One area where the current and UK studies differ is in the performance of the control toothpaste. In the current study, the control toothpaste showed significant improvements in gum health and plaque measures compared to baseline at all time points, whereas in the UK study, gingival and plaque scores increased from baseline [15]. The design of the studies differed, with participants in UK study having a professional prophylaxis and using the control toothpaste for 4 weeks before the baseline assessments, while participants in current study started using the study toothpastes immediately without any prior prophylaxis and run-in phase [15]. The UK design was employed to try and reduce the Hawthorne effect, whereby the gum health of those using the control toothpaste improves due to participants improving their brushing technique as a result of participating in the study [27, 15]. A comparison between control toothpaste data from the two studies would indicate that the approach used in UK study had the desired effect of reducing the Hawthorne effect while still demonstrating the benefit of the enzyme and protein containing toothpaste [15].

The test toothpaste incorporates a three enzyme system designed to naturally augment saliva’s natural defence systems. Amyloglucosidase and glucose oxidase combine to generate hydrogen peroxide from polyglucans, and then hydrogen peroxide is utilised by lactoperoxidase to convert salivary thiocyanate into hypothiocyanite [14,15]. Both hydrogen peroxide and hypothiocyanite are known for their antimicrobial action and have been demonstrated to inhibit bacteria involved in periodontitis [14,16,17]. This proposed mode of action has been supported through a series of *in vitro* studies which demonstrated that the test toothpaste increased levels of hypothiocyanite and hydrogen peroxide in saliva and provided antimicrobial effects in both single species and multispecies biofilm models [21]. Furthermore, twice daily brushing with the test toothpaste for 14 weeks was shown in a previous clinical study to shift the oral microbiome towards health [16]. This shift resulted from increases in relative abundance of bacteria associated with gum health, such as *Neisseria spp.,* and decreases in relative abundance of periodontitis related species such as *Treponema* [16].

Dental plaque is the main trigger of gingivitis [6]. If left untreated, gingivitis can advance to periodontitis, which not only affects oral health but also has broader systemic implications [8,9,10,11]. Maintaining proper oral hygiene is essential for preventing and managing gingivitis by minimising plaque build-up [8]. The enzyme and protein containing toothpaste demonstrates the ability to reduce gingival inflammation, both through reducing levels of dental plaque in the mouth and encouraging a healthier plaque microbiome. Hence, it has the potential to prevent gingivitis, retaining a strong connection between the gum and tooth, and helping to prevent the concomitant progression to periodontitis.

## Conclusion

In conclusion, this study demonstrated that brushing with toothpaste containing natural enzymes and proteins can significantly improve gingival health and reduce supragingival plaque levels among Chinese individuals with gum problems, compared to control fluoride toothpaste.

## Data Availability

The datasets generated and/or analysed during the current study are not publicly available as consent was not received from participants for this purpose.

## List of Abbreviations

AE: Adverse Event
ANCOVA: Analysis of Covariance
CI: Confidence Interval
CONSORT: Consolidated Standards of Reporting Trials
F: Fluoride
GI: Gingival Index
MQHPI: Modified Quigley and Hein Plaque Index
P: Percentage of sites with no bleeding
ppm: parts per million
SD: Standard Deviation
UK: United Kingdom

## Declarations

### Ethics approval and consent to participate

The study was performed in accordance with Good Clinical Practice, the Declaration of Helsinki and local legal and regulatory requirements. The study was approved by the ethics committee of the Hospital of Stomatology, Xi’an Jiaotong University. All participants provided written informed consent to participant in the study.

### Consent for publication

Not applicable.

## Author’s contributions

All authors have read and approved the manuscript. J.N.S. and J.R.M. contributed to the design of the study and the writing of the protocol and review. P.N.Z. led the study conduct. R.H. and J.T. led the study organization and supervision. X.H. contributed to original draft. J.R.M., L.Z., S.L. reviewed, edited and commented on manuscript drafts. A.K.G. was statistician for the study.

## Acknowledgements

The authors would like to thank stats team from Cytel Statistical Software & Services Pvt.Ltd India for conducting the statistical analysis, and Sinead Malone, Robert Marriott and Renuka V from Unilever for their valuable comments on manuscript drafts.

